# Intensity and appreciation of sweet taste solutions are modulated by high-intensity aerobic exercise in adolescent athletic males

**DOI:** 10.1101/2022.10.02.22280612

**Authors:** Alexandre-Charles Gauthier, Marc-Étienne Villeneuve, Mathieu Cournoyer, Marie-Eve Mathieu

## Abstract

**Introduction:** Exercise tends to reduce subsequent meal intake, but mechanisms are still unclear. Interestingly, exercise seems to influence taste, which plays a role in energy intake. The effect of exercise on specific tastes is still to be elucidated, especially among younger participants who train at high intensity.

**Methods:** Adolescents (14-16 years old) were recruited from a high school boys hockey team. Distinct taste tests were administered using low and high concentrations of sweet (sucrose 41.0 & 82.0 g/L), salty (sodium chloride 8.7 & 17.4 g/L) and bitter (caffeine 5.0 & 10.0 g/L) solutions before and after a 30 min aerobic high-intensity exercise session (70-90% of estimated maximal heart rate). McNemmar’s tests, standard paired T tests, Wilcoxon Signed Rank Test and Cohen’s d effect size tests were used to analyze the data.

**Results:** Participants (n=19) were 14.7±0.7 years old, weighed 59.6±7.8kg, had a height of 173.4±7.9cm, and a bodyfat% of 11.6±3.1%. There were no significant differences in taste identification capacities. Participants (n=19) perceived as more intense (+31%, p=0.037) and appreciated better the low concentration sweet solution (+20%, p=0.004). Taste appreciation was also increased for the high concentration sweet solution (+15%, p=0.009). Effect sizes were medium [0.516-0.776].

**Conclusion:** High-intensity exercise influenced the perception of sweet taste. If higher taste intensity and appreciation of sweet can reduce energy intake, our results could help explain the effect of exercise on lowering subsequent energy intake.

## 1. Introduction

Physical activity (PA) levels, which correspondingly induce energy expenditure, are partly responsible for energy intake (Bosy-Westphal, Hägele, & Müller, 2021). People who are more active tend to be more sensitive to their energy status. Thus, they either increase or decrease their energy intake in order to match their demand according to their appetite/satiety signals (Dorling et al., 2018). This control over their energy intake could play a positive role in attaining an energy-neutral state (Beaulieu, Hopkins, Blundell, & Finlayson, 2016). Even though different PA levels greatly dictate energy expenditure related to exercise, people who practice aerobic exercise do not necessarily increase their energy intake sufficiently to match the intensities and duration of the exercise (Donnelly et al., 2014; Taylor, Keating, Holland, Coombes, & Leveritt, 2018). Although acute exercise has a relatively small effect on energy consumption, active people tend to have lower body weight and more regulated appetite signals than inactive people. In fact, when comparing groups of people over one year, the odds of gaining more than 3% of fat mass were 1.8 to 3.8 times higher in the lower physically active group vs the physically active one (Shook et al., 2015).

While exercise has a definitive impact on subsequent feeding, the mechanisms behind this effect remain unclear. Indeed, feeding habits are closely related to energy homeostasis, environment, physiology, and emotions, all playing crucial roles in energy intake (Farr, Chiang-shan, & Mantzoros, 2016; Robinson, Thomas, Aveyard, & Higgs, 2014). The sensory system, including taste, also plays an important role in food intake, satiety signals and nutrient sensing (Boesveldt & de Graaf, 2017; de Graaf, 2020). Taste perceptions can also modulate food preferences, which could influence energy intake (Liem & Russell, 2019). Since exercise has a documented effect on nutrition and energy intake, it has been wondered whether taste could be one of the key explaining factors. Our lab recently conducted a systematic review study on the overall impact of exercise on taste perceptions (Gauthier et al., 2020). In this review, it was mentioned that exercise did increase sweet taste and salty taste preferences. While sensitivity and intensity of sweet taste were increased following exercise, they were both lowered for salty taste (Gauthier et al., 2020).

It is important to note that this systematic review included only 18 studies (Gauthier et al., 2020) and that although the impact of acute exercise and PA on taste has been documented in the past decades, the literature is still lacking overall content and knowledge regarding the matter. For example, the literature is still ambiguous regarding the impact of exercise on sweet taste appreciation. While appreciation was increased following acute exercise (T. Horio & Kawamura, 1998; T. J. P. Horio & skills, 2004; King, Appleton, Rogers, Blundell, & behavior, 1999; D. H. Passe, Horn, Stofan, Murray, & metabolism, 2004), people who practice regular PA have an overall lowered preference for sweetness (Crystal, Frye, & Kanarek, 1995; Feeney et al., 2019). To our knowledge, no study has investigated the impact of exercise on taste in younger populations (i.e. < 18 years old). Another important observation in our systematic review is that the studies included had different forms of exercise, where higher exercise intensities were less common (Gauthier et al., 2020). Most of them could be considered aerobic exercise, usually at lower intensities and maintaining a steady pace. The exercise sessions mostly contained submaximal exercise prescribed at a percentage of the VO2MAX (∼70%) or of the maximal heart rate (50 to 75% of HRmax) of the participants (Gauthier et al., 2020). While some examples of higher exercise intensities have been reported across the literature, they are usually done at a steady pace at around 80% of the participant’s HRmax (Havermans, Salvy, & Jansen, 2009; T. J. P. Horio & skills, 2004). To our knowledge, no study has examined the effect of higher exercise intensities on taste, potentially because sustained higher intensities are usually harder to perform for the general population. Since the literature is mainly done on healthy normal-weight individuals with some studies on athletic population (13), it was wondered whether the same results could be replicated in a young athletic adolescent population. Also, most of the literature regarding the impact of physical exercise on taste perceptions has documented sweet and salty tastes as outcomes (Gauthier et al., 2020). Bitterness is one of the least explored tastes, hence why it was essential to include this taste in the current study. While the literature on bitterness in relation to food consumption is still scarce, there is some evidence that bitter sensitivity and overall taste function could have implications regarding energy consumption (Choi, 2019; Graham et al., 2021).

Therefore, the objective of the current study was to investigate if athletic adolescent boys had better identification capacities and increased taste intensity perception and appreciation for individual tastes (sweet, salty and bitter solutions) following an acute session of ramping intensity aerobic exercise session. When compared to the pre-testing results, it was hypothesized that teenager athletes would have overall better identification capacities and that there would be a significant increase in their ratings of taste intensity for every solution, while taste appreciation of sweet and salty solutions would also increase following acute exercise.

## 2. Method

### 2.1. Study design and study population

Participants were recruited from a boys’ hockey team of a local high school, Collège Durocher, Saint-Lambert, Canada. Participants were between 14 and 16 years old, and all had been practicing with the team for at least two months. Exclusion criteria included dietary restraints, medication that could possibly alter taste perceptions, lasting COVID-19 side effects and/or if the participants a) had COVID-19 in the last six months and/or b) had physical exercise restrictions such as injuries or medical conditions at the time they signed up and participated in the study. They were asked not to do any exercise and not to drink any alcohol nor consume any drugs or tobacco for 12 hours prior to testing. Participants arrived at 9:00 a.m. and were separated into five different groups and were each given a letter corresponding to a randomised taste sequence. This research project was conducted at the Université de Montréal, precisely at the CEPSUM, and was approved by the ethics committee of health sciences (CÉRES) of the Université de Montréal (# 2021-392).

### 2.2. Experimental protocol

Each group was tested individually. Participants were first asked to consume an appetite normalizing snack, which consisted of ad libitum *Clif*TM bars, to reach an appetite feeling of the equivalent of four out of five 30 minutes before the first taste test. After the snack, their body weight and body fat mass were measured using bioelectrical impedance (TB350, Tanita, Arlington Heights, United States of America). Their height was measured with a portable stadiometer (SECA 217, Hamburg, Germany). Participants were then tasked to the French version of the International Physical Activity Questionnaire-Adolescent to assess their general PA levels (Hagströmer et al., 2008). Then, each participant underwent the taste evaluation sequence described below before moving on to the aerobic exercise session. The research team monitored each participant’s water consumption from the end of the first taste test until the last weight measurement. After the exercise session, each participant was asked to complete the same taste evaluation, which was randomised between participants in each group, and participants were weighed one last time before leaving (Figure 1).

**Figure 1.**
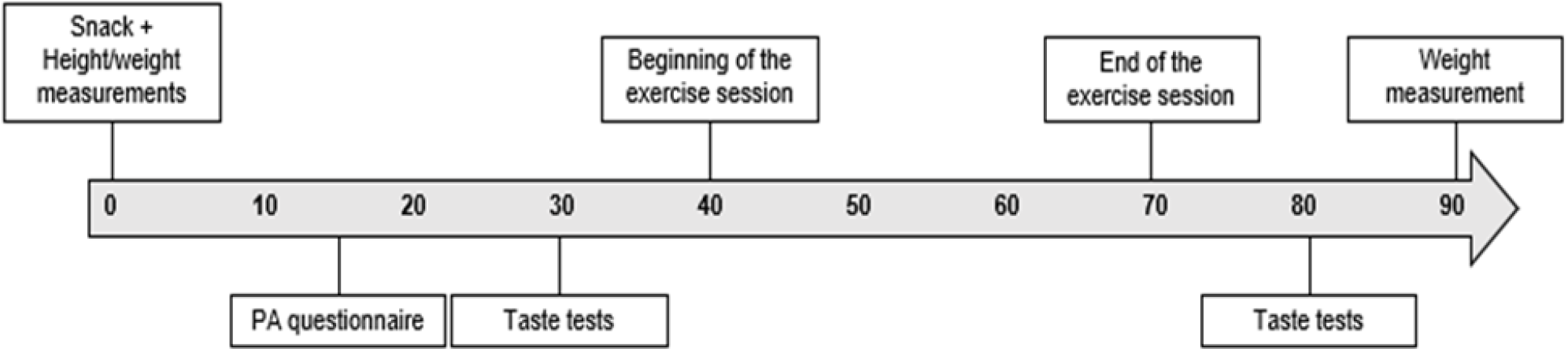
Testing sequence in minutes for each participant.

**Figure 2.**
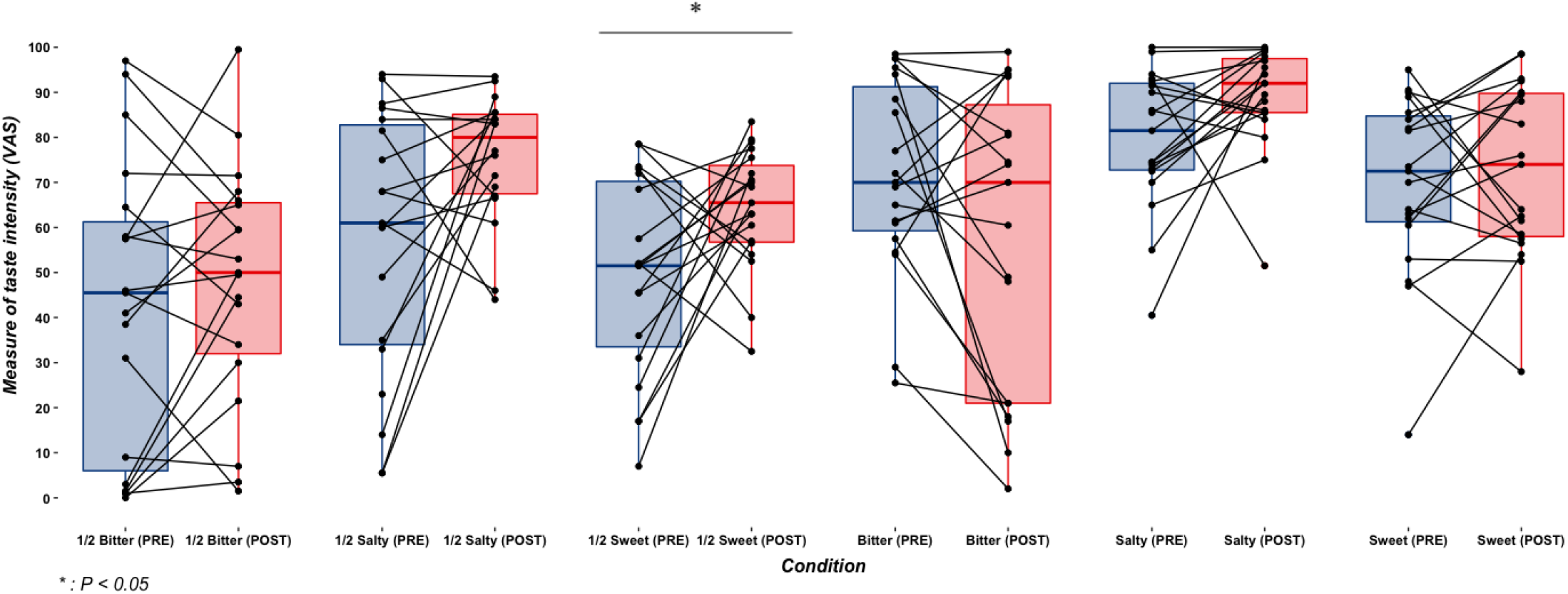
Salty and Sweet taste intensity before and after exercise. Box plot represents the median with the lower and upper quartile. The vertical lines represent the minimum and the maximum.

**Figure 3.**
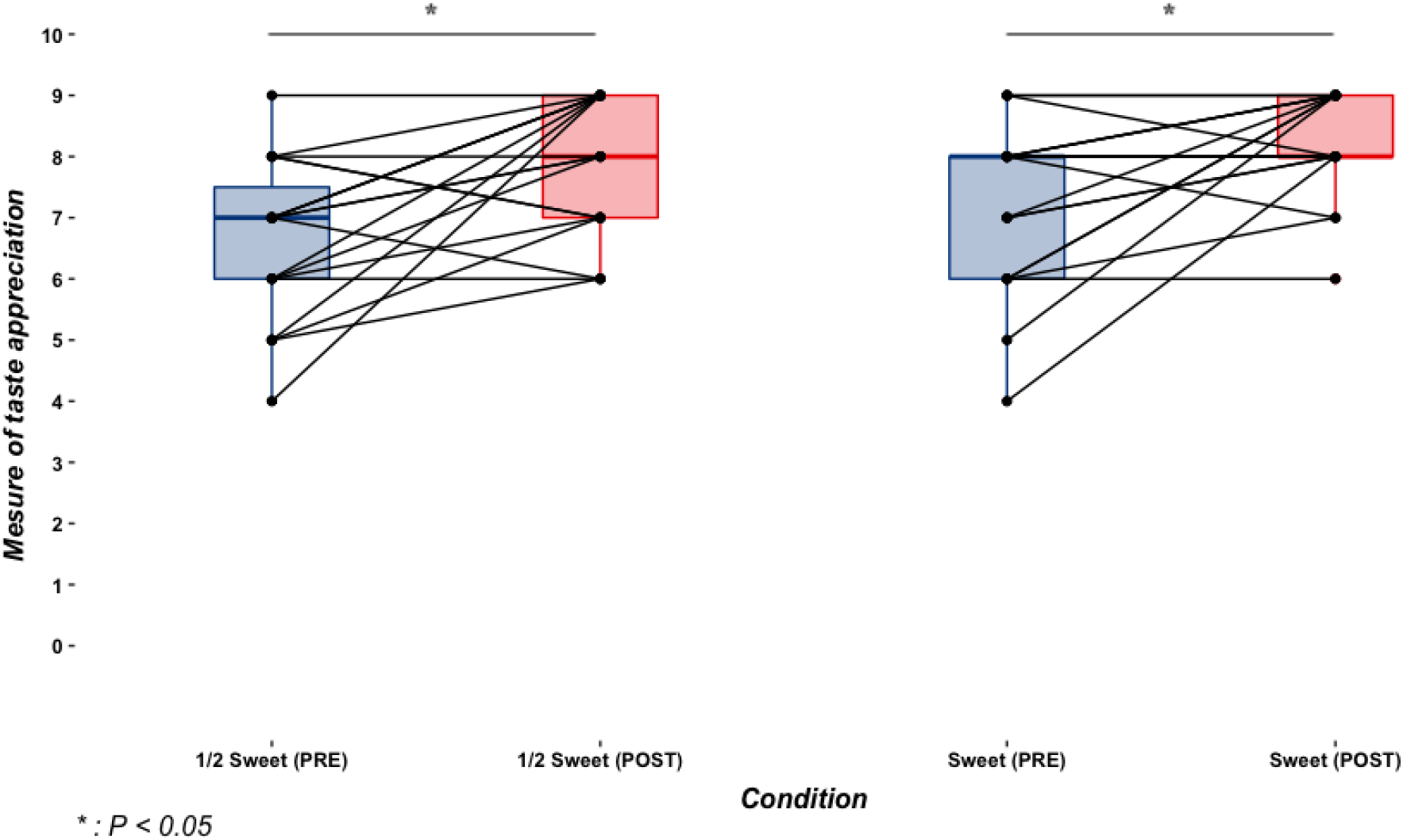
Sweet taste appreciation before and after exercise. Box plot represents the median with the lower and upper quartile. The vertical lines represent the minimum and the maximum.

#### Taste test protocol

Each individual was presented with the same randomised cup order before and after exercise. Disposable cups were filled with four different tastes: sweet, salty, bitter and nothing (water). Within the different tastes, solutions were divided into three categories: half concentrated, fully concentrated and water. The solutions were presented under red lights, so they could not discern each solution’s colour. Each participant had to identify seven solutions individually, and they also had to evaluate the perceived intensity and appreciation of the solution. For the identification question, participants were given five choices: bitter, sweet, salty, no taste and other. Bitter solutions had a concentration of 10.0 g/L and 5.0 g/L (caffeine pills), respectively, sweet solutions had a concentration of 82.0 g/L and 41.0 g/L (sucrose), respectively, and salty solutions had a concentration of 17.4 g/L and 8.7 g/L (sodium chloride) respectively.

Intensity ratings were done on a 0 to 100 mm visual analog scale (VAS), ranging from no taste whatsoever to the strongest taste experienced. The usage of VAS has been one of the cornerstones of sensory evaluation as it seems to offer precise subjective results and a precise tool to offer evidence of treatment (Bijur, Silver, & Gallagher, 2001). The appreciation scale was a standardised 9-point hedonic scale on which participants had to give a score ranging from 1 (dislike extremely) to 9 (like extremely), 5 being neither like nor dislike for every solution. The 9-point hedonic scale is a standardised scale which has been validated and is used to assess food/taste preferences (Stone & Sidel, 2004). Each participant was assigned to an individual table, so they would not be able to see the other participants. Participants were asked to rinse their mouths between each taste solution by swirling back and forth water in their mouths eight times.

### 2.3. Exercise protocol

Each participant wore a heart rate monitor (Polar Team2 Pro, Kempele, Finland) for the exercise session. Participants were first asked to jog lightly for 3 to 5 minutes around the running track to warm up. Then, the exercise session had participants track running for 30 minutes with increasing intensity every 10 minutes. They all started at 70% of their HRmax for 10 minutes, then 80% of the HRmax for 10 minutes, and then 90% of their HRmax for 10 minutes. HRmax, according to a recent systematic review on the prediction of the HRmax of children and adolescents, was set at 197 beats per minute (Cicone et al., 2019; Gelbart, Ziv-Baran, Williams, Yarom, & Dubnov-Raz, 2017). Participants had to be within five beats per minute of their targeted heart rate for the intensity that was indicated, and a researcher monitored (Polar Team2 Pro, Kempele, Finland) each group for the whole duration of the exercise session.

### 2.4. Statistical analyses

Age (years old), height (cm), body mass (kg), body fat percentage (%) and water consumption (mL) were presented as a mean ± standard deviation. McNemmar’s tests, standard paired T tests, Wilcoxon Signed Rank Test and Cohen’s d effect size tests were performed to analyze the data regarding differences in means for pre and post exercise measurements for weight, taste identification test, taste intensity test, and taste appreciation test using IBM SPSS Statistics 27TM (Endicott, United States of America). Cohen’s d classification scale is as follows: very small (0 ≤ Cohen’s d <0.20), small (0.20 ≤ Cohen’s d <0.50), medium (0.50 ≤ Cohen’s d <0.80), and large (Cohen’s d ≥ 0.80).

## 3. Results

### 3.1. Population characteristics

Participants (n=19) had a mean age of 14.7±0.7 years, a mean height of 173.4±7.9 cm, a mean body mass of 59.6±7.8 kg and a mean body fat percentage of 11.6±3.1 %. They consumed on average 298.7±142.8 ml of water during the experiment. Significant differences were obtained when comparing pre and post exercise body mass with an average loss of 0.4 kg difference (p<0.001). Participants we’re all classified as high level of PA according to their METS-minutes/week results for the IPAQ-A.

**Table 1.** Participants general characteristics.

| | mean ± SD |
| --- | --- |
| Age | 14.7 ± 0.7 |
| Body Weight |  |
| PRE | 59.7 ± 7.8 |
| POST | 59.3 ± 7.8 |
| % Body Fat |  |
| PRE | 11.6 ± 3.1 |
| POST | 9.5 ± 3.0* |
| Height | 173.4 ± 8.0 |
| Water Consumption | 298.7 ± 142.8 |
| METS*min/Week | 5429.3 ± 1919.3 |
| * $P = .041$ | |

### 3.2. Taste test results

#### Identification test

To analyse the intensity data, McNemar’s tests were used for every solution. No significant results were obtained regarding the identification of each solution.

**Table 2.** Identification test results. Results are displayed as a percentage of rightfully identified solution.

| Taste | Pre | Post | p-value |
| --- | --- | --- | --- |
| **Bitter** | 67% | 72% | 0.564 |
| **½ Bitter** | 63% | 79% | 0.257 |
| **Salty** | 89% | 94% | 0.317 |
| **½ Salty** | 83% | 83% | 1.000 |
| **Sweet** | 100% | 100% | 1.000 |
| **½ Sweet** | 100% | 90% | 0.157 |
| **Water** | 83% | 83% | 1.000 |

#### Intensity test

To analyse the intensity data, standard paired T tests were used for every solution. Only the half concentrated sweet solution for the intensity test yielded significant results with a mean pre score of 49.1/100 mm and a post score 64.3/100 mm (p=0.037, d=0.516).

While other taste solutions such as half and fully concentrated salty solutions did not reach significance, they were almost classified as medium effect sizes (p=0.0501, d=0.497 and p=0.0543, d=0.472, respectively). Half concentrated bitter solutions (p=0.348, d=0.221), and fully concentrated bitter (p=0.084, d=0.420) and sweet (p=0.570, d=0.133) solutions did not change significantly and were all classified as very small or small effect size according to Cohen’s d parameters.

#### Appreciation test

To analyse the appreciation data, Wilcoxon Signed Rank tests were used for every solution. Both half and fully concentrated sweet solutions for the appreciation tests increased significantly: the half concentrated sweet taste solution had a mean pre score of 6.6/9.0 and a post score 7.9/9.0 (p=0.004, d=0.776) and the fully concentrated sweet taste solution had a mean pre score of 7.2/9.0 and a post score 8.3/9.0 (p=0.009, d=0.698).

Half (p=0.578; d=0.130) and fully concentrated (p=0.164; d=0.341) salty solutions as well as half (p=0.447; d=0.147) and fully (p=1.000; d=0.000) concentrated bitter solutions did not change significantly, and their effect sizes were considered very small or small.

## 4. Discussion

The objective of the current study was to investigate if athletic adolescent boys had better identification capacities and increased taste intensity and appreciation for distinct sweet, salty and bitter solutions following an acute session of ramping intensity aerobic exercise. This project has the unique characteristic of studying the evolution of individual taste perceptions in young males who exercise almost daily in response to an exercise session. To our knowledge, it is also the first study with a ramping high-intensity running session as its exercise component in a younger population compared to other protocols in the exercise-taste literature. While identification did not differ from pre- to post-exercise session, taste appreciation was augmented for half and fully concentrated sweet solutions. Intensity for the half concentrated sweet solution was also increased following exercise, but the fully concentrated sweet solution offered no such results. While some trends were present for the half-concentrated and fully concentrated salty solutions, especially regarding their intensity, no significant results were obtained. There were no significant results for the half concentrated and fully concentrated bitter solutions for the identification, intensity, and appreciation tests.

### Sweet

No significant results were obtained for the identification taste test for the sweet solutions. Even though each half-concentrated solution had a subtle taste, participants could clearly distinguish each taste individually. Conventionally, this kind of testing can be done in a sensitivity testing fashion. Taste sensitivity testing is used to assess the minimum threshold at which a person can detect a specific taste and identify it (Sørensen, Møller, Flint, Martens, & Raben, 2003). This kind of testing usually involves different concentrations of the same tastant, which can be presented in a randomised order. Since this protocol had a time constraint, it was only feasible to fit two concentrations per taste for the identification test.

People who exercise regularly tend to rate sweet taste intensity higher when compared to controlled groups (Crystal et al., 1995; Feeney et al., 2019; Kanarek, Ryu, Przypek, & behavior, 1995). Furthermore, as reported by Ali and al., intensity ratings for sweetness are at their highest during the exercise session compared to pre and post-conditions (Ali et al., 2011). While our protocol did also report some significant increases in intensity ratings regarding sweet taste, it was only with the half-concentrated sweet solution. For the half concentrated sweet taste solution, a significant increase of ∼31% was obtained when comparing post-exercise values to pre-exercise values with an effect size that would be considered medium, with a score of 0.516 under the Cohen’s d parameters. One fundamental explanation of these results could be that the fully concentrated solution already had high intensity ratings at baseline when compared to the half-concentrated solution, suggesting a ceiling effect. The difference in mean ratings between the two solutions at baseline was 20 mm on the 100 mm scale, with values hovering around 49.0 mm and 69.7 mm, respectively. The latter result is a byproduct of the already highly concentrated solution. Feeney et al., who used roughly the same concentration as in this protocol for the higher concentration sucrose solutions (∼82.0 g/L) to evaluate sweetness intensity, found a significant difference between active and inactive populations (Feeney et al., 2019). While these results differ from ours, the participants in our current study were all active individuals and taste tests were obtained before and after a single acute exercise session compared to Feeney’s protocol, where exercise levels were self-reported, giving an overall view of their PA levels. Narukawa and al., who used even higher concentration solutions for the highly concentrated sucrose solution (∼102.0 g/L) with a much longer exercise session (36 km mountain hike for 12 hours), also found no significant difference between pre- and post-exercise acute results (Narukawa et al., 2010), which is somewhat in agreement with the current study.

When looking at the literature, the vast majority is done on appreciation parameters, such as perceived pleasantness, preferences, hedonic ratings and overall liking (Gauthier et al., 2020). Reports of increased preference for sweetness following exercise were done with varying concentrations of sucrose (T. Horio & Kawamura, 1998; T. J. P. Horio & skills, 2004) or with sweet fruit drinks (King et al., 1999). While King and al. used visual analog scales to assess sweet preferences (King et al., 1999), Horio and Kawamura used a 7-point hedonic scale (T. Horio & Kawamura, 1998), and Horio used a 9-point hedonic scale (T. J. P. Horio & skills, 2004). Our protocol also used a hedonic 9-point evaluation scale to analyse taste appreciation pre- and post-exercise. In line with the aforementioned literature, the current study did report significant increases of ∼20% and ∼15% for sweet taste appreciations for the half and fully concentrated sweet solutions, respectively, following acute physical exercise. The effect size for both half and fully concentrated sweet taste solutions would be considered medium, with a d=0.776 and d=0.698, respectively. Under Cohen’s d parameters, the half-concentrated sweet solution is on the edge of a large effect size (Cohen’s d ≥ 0.80). These results add compelling evidence that sweet preference is increased following an acute bout of exercise.

### Salty

For the identification test of the salty solutions, participants were mostly able to quickly identify each salty solution, both for the half and the fully concentrated ones. Similarly to the sweet taste identification, no significant results were obtained. This could partly be explained by the fact that the salty solutions were also heavily distinguishable from the other taste solutions at baseline (pre=84.2% V.S. post=83.3%, p>0.05 and pre=88.9% V.S. post=94.7%, p>0.05, for half and fully concentrated solutions respectively).

A significant decrease in saltiness intensity has only been reported in another study (Ali et al., 2011). Ali and al. conducted a study in which participants’ intensity ratings were lower during the exercise session compared to the pre-conditions (Ali et al., 2011). Also, the duration of the exercise had a significant impact on these intensity ratings, with differences from pre- (30 min before) to post-exercise ranging from ∼31% to ∼48% difference for the higher electrolyte-containing solutions [2] (Ali et al., 2011). While the results obtained in the current study did not reach significance, participants rated the half-concentrated salty solution with a ∼22% increase in intensity scores following the exercise session compared to the pre-exercise score (p=0.0501). Similarly, the intensity of the fully concentrated salty solution was ∼11% higher when compared to the pre-exercise session (p=0.0543). Our base ratings for the half and fully concentrated salty solutions were at 58.9 mm and 79.6 mm, with concentrations of 8.7 g/L and 17.4 g/L of sodium chloride, respectively. To put in perspective, Ali and al. had base saltiness ratings for the higher electrolyte solutions [2] on the visual analog scales of 32.6 mm and 34.6 mm out of 100 mm, with these solutions containing 0.280 g/L of sodium and 0.141 g/L of potassium (Ali et al., 2011). High baseline intensity ratings could partly explain why no significant differences were present for both salty solutions in the present study. The effect sizes for both half and fully concentrated salty solutions (d=0.497 and d=0.472, respectively) could almost be considered as a medium effect sizes. Considering these effect sizes, it could be interesting to keep exploring the link between salt taste intensity and acute physical exercise since a medium effect size is non-negligible. With these results, we could speculate that lower baseline ratings of saltiness combined with more participants could have yielded significant findings and bigger effect sizes. Alas, they were not obtained with the current settings.

As previously mentioned, the bulk of the literature is concentrated around preferences and appreciation for overall taste testing (Gauthier et al., 2020). Across all articles that reported changes in taste preference for saltiness in link with physical exercise, a significant increase was noted for acute exercise (Leshem, Abutbul, & Eilon, 1999; D. Passe, Stofan, Rowe, Horswill, & Murray, 2009; Wald & Leshem, 2003) and regular PA levels (Kanarek et al., 1995). This study did not provide significant results regarding increased appreciation for salty solutions. One explanation might be that the overall liking of the salt solutions was relatively low at the very start. Since these solutions were only composed of salt and water, it could explain why they might have been deemed unpleasant from the beginning. Some authors also reported no change following exercise in preference for saltiness (T. Horio & Kawamura, 1998). It is to be noted that in the literature when the preference for saltiness is tested following exercise, it is mostly combined with other tastes/ingredients such as sweet and/or sour taste in a solution (Ali et al., 2011; T. J. P. Horio & skills, 2004; D. Passe et al., 2009) or in certain foods that contain varying levels of salt (Leshem et al., 1999). Nevertheless, while the current results align with the overall literature on the matter, no significant differences were obtained while analysing the data.

### Bitter

Bitterness is the only one of the five primary tastes that has not produced any significant results in relation to acute exercise and PA in the literature despite being tested in 5 studies (Gauthier et al., 2020). While the bitter taste was included in our protocol with all three sensory taste tests, both as a half and a fully concentrated solution, it offered no significant results. Bitter is a taste which was primarily used to detect poisonous food up until recent centuries (Smail, 2019). We sought to see whether we could generate new data regarding the impact of physical exercise on bitterness, but none was obtained. As of now, it is safe to assume that acute exercise and PA might not have any impact on the bitter taste.

### Strength and limitations

Our study is one of the first to evaluate the impact of physical exercise on individual taste perceptions in a younger active population with a ramping exercise high-intensity protocol. Standardising the pre-testing conditions for every participant, especially regarding their appetite/hunger status and hydration status, was one of the main characteristics of this study. In other studies, most participants were asked to limit their PA levels the day before or the day of the testing and to not eat before testing, but their energy intake and their appetite were mostly not documented and/or standardised (T. Horio & Kawamura, 1998; T. J. P. Horio & skills, 2004; Narukawa et al., 2010). Since these two could play a potential role in our perception of taste and are influenced by the moment of the testing and our energy status (Nakamura et al., 2008), it is thus crucial to control these parameters to draw more robust conclusions. While our protocol only included a 30-minute exercise session, significant results arose for the sweet taste. A longer duration might have generated different results, especially regarding the salty taste, since longer exercise duration is usually associated with higher electrolyte depletion. However, athletes who did a 40-minute running session at a speed intensity 10% under their lactate threshold did not report any significant increase in their salt preference using visual analog scales (Manevitz et al., 2021). Although a longer period of exercise would have been ideal to examine this possibility, it was unfortunately impossible considering the ongoing COVID restrictions that were in place at the time of the testing day. Also, while we did manage to test 19 participants, a higher number of participants than was anticipated beforehand. When screening the literature, successful protocols had around 30 participants in their project. Meanwhile, we did not have any exclusion criteria based on the sex of the participant, this project only tested adolescent boys due to our participant pool. The differences between women and men regarding taste perceptions has yet to be studied. Moreover, sensitivity testing is one of the cornerstones of taste perception testing and is often used in the exercise-taste literature. The usage of more solutions with increasing concentrations would have made the identification test more prone to detect some sensitivity changes following acute exercise. The testing sequence revealed that it would not have been feasible with the participants’ time at hand and attention span. Hence, our paper failed to provide satisfactory results regarding sensitivity changes in link with exercise. Future studies that seek to increase overall knowledge regarding the impact of exercise on taste should include accurate sensitivity testing with appropriate solution concentrations and longer periods of exercise.

## 5. Conclusion

To summarize, this paper produced significant results regarding the impact of acute physical exercise on distinct taste perceptions in an active adolescent population. Appreciation scores and intensity scores for sweet tasting solutions were at the heart of the results. While the specific effects of acute exercise on energy intake still lack concrete pathways, taste could be part of the equation. While the link between exercise and taste has yet to be fully exploited, it would be interesting to analyse whether modulated taste intensities and appreciation of sweet have an influence over energy intake. This paper sought to deepen the understanding of this matter while providing new data to be compiled with the existing knowledge.

## Data Availability

All data produced in the present study are available upon reasonable request to the authors.

## Acknowledgement

A special thank you to David Ouellette, the kinesiologist and hockey coach who played a pivotal role in recruitment and the overall success of the testing sequence. ME Mathieu holds a Canada Research Chair – tier 2 on Physical activity and juvenile obesity that helped finance the project.

